# Clinical correlation of lung ultrasound profiles in patients with COVID-19 infection

**DOI:** 10.1101/2021.04.05.21254935

**Authors:** Chitrakshi Nagpal, Sanchit Kumar, Naveet Wig, Arvind Kumar, Praful Pandey, Manraj Singh, Kapil Dev Soni, Richa Aggarwal, Anjan Trikha

## Abstract

**Background:** Lung ultrasound is a popular point of care test that correlates well with computed tomography for lung pathologies. While previous studies have shown its ability to detect COVID-19 related lung pathology, we aimed to evaluate the utility of lung ultrasound in the triage and prognostication of COVID-19 patients by determining its ability to predict clinical severity and outcomes.

**Methods:** This was a prospective, cross-sectional, observational, single centre study done at JPNATC and AIIMS, New Delhi, India. Consenting eligible patients aged 18 years or more were included if hospitalised with microbiologically confirmed COVID-19 and classified as mild, moderate (respiratory rate >24/min OR SpO2<94% on room air) and severe COVID-19 (respiratory rate >30/min OR SpO2<90% on room air) at the time of enrolment. The lungs were systematically assessed with ultrasound after division into 14 zones (4 anteriorly, 4 axillary and 6 posteriorly). Clinical and laboratory parameters including arterial blood gas analysis at the time of evaluation were recorded. Patients were followed till death or discharge. The primary objective was to determine the correlation between clinical severity and lung ultrasound profiles (no. of A, B and C profiles, and the total number of areas involved). Secondary objectives included assessment of the correlation between lung ultrasound profiles and clinical outcomes and development of a statistical model incorporating ultrasound and clinical parameters to allow prediction of COVID-19 related severity and outcomes.

**Findings:** Between October 1, 2020, and January 31,2021, patients were screened for inclusion and total n=60 patients were evaluated and included in the final analysis. The most common abnormality seen were B lines, seen in at least one zone in n=53 (88.33%) of cases. A median of 9 (IQR: 5-12) zones of the 14 assessed had a B-profile. The total number of abnormal areas (zones with a B or C profile) correlated significantly with the PaO2/FiO2 ratio (ρ= −0.7232, p<0.0001) and SpO2/FiO2 ratio (ρ= −0.6866, p<0.0001), and differed significantly between mild and moderate vs severe cases (p=0.0026 mild vs moderate, p<0.0001 mild vs severe, p=0.0175 moderate vs severe). The total number of B lines were predictors of mortality (p=0.0188, OR 1.03, 95% CI 1.003-1.060). Statistical models that incorporated total number of B-lines, CRP and anticoagulation use could predict mortality (p=0.0124, pseudo R2=0.1740) with an AUC= 0.7682 (95% CI=0.6176-0.9188), and the total number of involved areas and LDH levels could distinguish severe disease from mild/moderate disease (p<0.0001, Pseudo R2=0.3822), AUC = 0.8743 (95% CI=0.7752-0.9733). A simplified cut off of ≥6 involved areas (of the 14 assessed) was 100% sensitive and 52% specific for differentiating severe disease from mild and moderate ones.

**Interpretation:** In patients with COVID-19, increasing involvement of the lungs as assessed by ultrasonography correlates significantly with clinical severity and outcomes. These findings may be utilized in future prospective studies to validate the use of lung ultrasound to triage and prognosticate patients with COVID-19 infection.

**Author Approval:** All authors have seen and approved the manuscript

**Competing interests:** There are no potential competing interests

**Data availability Statement:** All data referred to in the manuscript shall be provided when asked for.

**Disclaimers:** The authors have nothing to disclose

**Funding statement:** No funding source was involved.

## Introduction

As of 17^th^ February 2021, there have been 109,068,745 confirmed cases of COVID-19, including 2,409,011 deaths, reported by WHO^1^ with India accounting for 10%^2^. This pandemic has created an urgent need for the development of modalities that might help with the diagnosis, triaging, and appropriate management of the affected individuals.

Computed tomography (CT) of the chest has been the radiological modality of choice for evaluation of COVID-19 related lung pathology - showing bilateral, subpleural predominant ground glass opacities (GGOs) in the majority of cases, with validated scores for reporting and grading the severity of infection.^3^ However, apart from radiation exposure, issues pertaining to availability, logistics, manpower and adequate infection control make CT imaging unsuitable for triaging or continuous monitoring of large patient populations.

Bedside lung ultrasound provides several benefits over CT imaging including portability, inexpensive testing, lack of radiation and instantaneous image generation. Studies on the use of lung ultrasound in COVID-19 infection showed that almost all patients affected by SARS-CoV-2 had abnormalities that could be detected using lung ultrasound, most commonly as a B profile suggestive of interstitial involvement^4^. Other findings in decreasing order of prevalence included pleural line abnormalities, pleural thickening, subpleural or pulmonary consolidation and pleural effusions in decreasing order of frequency. Good corroboration was noted between chest CT findings and bedside lung ultrasound, including severity scores.^5^

However, data on relationship between lung ultrasound findings and clinical parameters remains scarce. We therefore conducted this study to correlate the severity of COVID-19 infection and subsequent outcomes with the abnormalities seen on lung ultrasonography.

## Materials and Methods

This was a prospective, cross-sectional, observational, single centre study done at JPNATC and AIIMS, New Delhi between October 1, 2020 and January 31, 2021. The study was reviewed and cleared by the Institute Ethics Committee (IEC). Informed consent was obtained from the participants/next of kin prior to enrolment.

Patients were eligible for inclusion if aged ≥ 18 years and admitted to our centre with a diagnosis of COVID-19 confirmed by Reverse-Transcriptase Polymerase Chain Reaction, TruNAAT, Cassette Based Nucleic Acid Amplification Test or rapid antigen test for SARS-CoV-2 in appropriate respiratory tract samples. Participants were classified as - moderate COVID-19 (respiratory rate of ≥ 24/min OR SpO2 ≤ 94% on room air) and severe COVID-19 (respiratory rate ≥30/min OR SpO2 ≤90% on room air OR the need for invasive or non-invasive mechanical ventilation) on enrolment. Initial protocol was later amended and approved by the IEC to also include mild cases (not fulfilling criteria for moderate or severe COVID-19) as well. Sample size of convenience – n=60 patients was taken.

Exclusion criteria included lack of consent, clinical records showing alternate pulmonary pathologies including hospital/ventilator acquired pneumonia, pneumothorax, moderate to large pleural effusion, interstitial lung disease, uncontrolled asthma or chronic obstructive pulmonary disease, lung abscess, acute decompensated heart failure, fluid overload, bronchiectasis or lung malignancy.

The primary objective was to determine the correlation between clinical severity and lung ultrasound profiles (no. of A, B and C profiles, and the total number of areas involved). Secondary objectives included assessment of the correlation between lung ultrasound profiles and clinical outcomes and development of a statistical model incorporating ultrasound and clinical parameters to allow prediction of COVID-19 related severity and outcomes.

Demographic details, comorbidities, days from symptom onset, baseline clinical parameters (blood pressure, heart rate, respiratory rate, Glasgow Coma Scale, SpO2), mode of oxygen delivery (interface, FiO2, ventilator mode and settings), baseline laboratory parameters including a complete blood panel (CBC), renal and liver function tests (RFT and LFT), and inflammatory markers including interleukin-6 (IL-6), procalcitonin, lactate dehydrogenase (LDH), C-reactive protein (CRP) and ferritin levels (if available), arterial blood gas (ABG) analysis and ongoing medications at the time of assessment were recorded. For patients not receiving oxygen by a fixed FiO_2_ device, FiO_2_ was calculated as 21% + 3 x oxygen flow in L/min. Follow-up data on outcomes (death or discharge) was obtained after a maximum of 28 days after enrolment using hospital patient records.

Lung USG was performed by the authors (CN, SK) who had received lung ultrasonography training as a part of the residency curriculum. The examination was performed over both lungs by dividing the chest into 14 regions-4 anteriorly, 4 in the axillary areas and 6 posteriorly (*Figure 1S)*. The examination as carried out using the curvilinear probe on the M-Turbo® ultrasound system (*FUJIFILM Sonosite Inc*.) held in a longitudinal axis, perpendicular to the ribs, and the observed profiles (*Figure 2S)* were recorded. Profiles were defined for individual zones as per existing literature.^6,7^ The investigators were blind to any other imaging performed on the patients.

Analysis was performed using STATA v12.0. Categorical variables were described by frequency tables. Continuous variables were analysed for normal distribution using Shapiro Wilk’s test and expressed as medians with interquartile ranges. Categorical variables were tested for significance using Chi-square or Fischer’s exact test. Mann Whitney Utest and Kruskall-Wallis (followed by Dunn’s test with Benjamini-Hochberg adjustment if significant) were applied for testing differences in continuous outcomes of nominal or ordinal variables. Correlation between continuous variables was examined using Spearman’s rank correlation coefficient. Logistic regression with analysis of maximum likelihood and odds ratio estimates was applied to test continuous variables (involved areas on the lung ultrasound) for binary outcomes (death vs discharge and mild-moderate vs severe). Variables with p<0.2 on univariate logistic regression for outcomes/severity were included in the multivariable logistic regression model and ROC curves were obtained to determine optimal cut-offs for prediction of severity and outcomes. A value of p<0.05 was considered significant.

## Results

A total of n=72 patients that met the inclusion criteria were screened for enrolment from October 1, 2020 till January 31, 2021. Of these, n=60 patients were finally included in the analysis.

**Figure 1.**
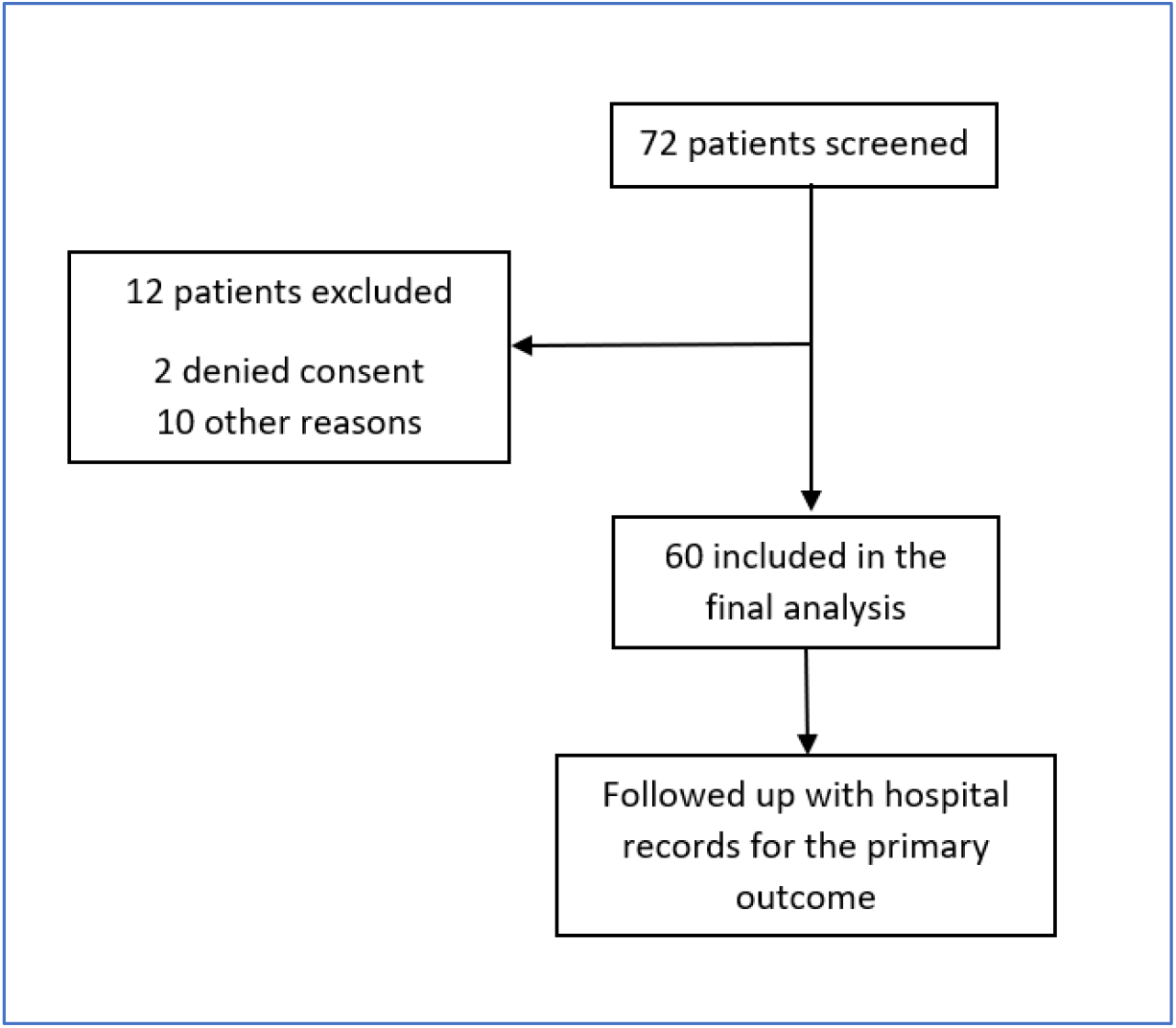
Flow chart of the participants’ recruitment process for LUS examination

*Table 1* shows the baseline characteristics of included patients. Of these, n=40 (66.67%) were males, with a median age of 60 (IQR 43.75-68.50) years. The most common comorbidity was hypertension, n=33 (55%). The scans were done after a median of 4 (2-8.25) days from symptom onset. At the time of evaluation, n=13 (21.77%) patients were receiving invasive mechanical ventilation (IMV), n=40 (66.67%) were receiving supplemental oxygen in other forms and n=10 (16.7%) required vasopressor support. The laboratory parameters and treatment received till the time of recruitment are summarised in *Table 2*. On follow up, n= 34 (56.7%) patients were discharged or transferred out after recovery from COVID-19 infection while n=26 (43.3%) patients died in hospital.

**Table 1:** Baseline characteristics.

| Baseline characteristics | median (IQR) |
| --- | --- |
| Age (years) | 60 (43.75-68.50) |
| Male gender, $n$ (%) | 40 (66.67%) |
| Days from symptom onset | 4 (2-8.25) |
| **Comorbidities:** | **N (%)** |
| Diabetes Mellitus | 22 (36.67%) |
| Hypertension | 33 (55%) |
| Hypothyroidism | 5 (8.33%) |
| Chronic Kidney disease | 14 (23.33%) |
| Chronic Obstructive Pulmonary Disease | 8 (13.33%) |
| Malignancy | 7 (11.67%) |
| Cerebrovascular accident | 8 (13.33%) |
| **Clinical Severity** | n (%) |
| Mild | 10 (16.67%) |
| Moderate | 15 (25%) |
| Severe | 35 (58.33%) |
| **Clinical Parameters** | median (IQR) |
| Respiratory rate (*breaths/min*) | 25 (21.5-30) |
| MAP (*mm Hg*) | 91.83 (83.58-100.5) |
| SpO2 % | 97 (94-100) |
| FiO2% | 55 (35.5-80) |
| SpO2/FiO2 ratio | 175.68 (125-274.60) |
| Vasopressor use, n (%) | 10 (16.67%) |
| **Oxygenation** | n (%) |
| Invasive Mechanical Ventilation | 14 (23.33%) |
| Non-Invasive Ventilation | 9 (15%) |
| High Flow Nasal Canula | 15 (25%) |
| Non-Rebreather Face Mask | 5 (8.33%) |
| Face Mask or Nasal prongs | 10 (16.67%) |
| Room air | 7 (11.67%) |

**Table 2.** Clinical characteristics and treatment.

|  |  |  |
| --- | --- | --- |
| **1.** | **Baseline laboratory parameters** | median (IQR) |
|  | Hb *gm/dL* | 10.85 (8.8-12.45) |
|  | TLC */uL* | 12900 (9970-15975) |
|  | Neutrophil count */uL* | 11231.50 (7184.5-14483.25) |
|  | Lymphocyte count */uL* | 1014.5 (737.75) |
|  | Platelets *million/uL* | 0.191 (0.147-0.286) |
|  | Urea *mg/dL* | 60 (38-133.5) |
|  | Creatinine *mg/dL* | 1 (0.7-2.2) |
|  | T. bilirubin *mg/dL* | 0.9 (0.6-1.4) |
|  | IL-6 *pg/mL* (n=51) | 28.17 (15.31-69.51) |
|  | CRP *mg/dL* (n=49) | 9.5 (5.04-17.55) |
|  | D-dimer (n=26) | 2.55 (0.6-7.99) |
|  | Ferritin *ng/mL* (n=54) | 622 (303-1170) |
|  | LDH *U/L* (n=53) | 384 (290-526) |
| 2. | **Arterial blood gas** |  |
|  | pH | 7.43 (7.39-7.47) |
|  | PaO2 *mm Hg* | 75 (65-111) |
|  | PaCO2 *mm Hg* | 33 (29-41) |
|  | HCO<sub>3</sub><sup>-</sup> *mmol/L* | 22 (18.25-26) |
|  | Lactate *mmol/L* | 1.3 (1-1.7) |
|  | PaO2/FiO2 Ratio | 157.58 (95.71-270) |
| 3. | **Treatment received** | n (%) |
|  | Steroids | 53 (88.33%) |
|  | Dexamethasone | 10 (16.67%) |
|  | Methyl Prednisone | 43 (71.66%) |
|  | Anticoagulants n (%) | 50 (83.33%) |
|  | High dose* | 41 (68.33%) |
|  | Low dose<sup>#</sup> | 9 (15%) |
|  | Remdesivir | 21 (35%) |
|  | Tocilizumab | 4 (6.67%) |
|  | Vit C and Zinc | 60 (100%) |
|  | Doxycycline | 57 (95%) |
|  | Ivermectin | 19 (31.67%) |
|  | Plasma therapy | 1 (1.67%) |

(\*High dose defined as Enoxaparin $\geq 40$ mg BD; Dalteparin $\geq 2500$ IU BD)
(# Low dose defined as Enoxaparin $\leq 60$ mg OD; Dalteparin $\leq 2500$ IU OD)

On lung ultrasound assessment, the most common abnormality found was a B profile with n=53 (88.33%) patients having at least one zone with a B profile. Of the 14 zones evaluated, median number of uninvolved zones (i.e., A profile suggestive of normal aeration) were 4 (range 0-14). The median number of involved areas (B/B’ or C/C’ profiles) were 10 (6-13), with a median of 9 (5-12) zones with B profiles and 2 (0-4.25) zones with C profiles. The ultrasound findings are summarised in *Table 3*. There was no difference in the involved areas between the anterior, middle and posterior chest (p=0.6592).

**Table 3.** Lung Ultrasound abnormalities.

| Lung ultrasound profile | Median (min-max) |
| --- | --- |
| A profile (no. of zones) | 4 (0-14) |
| B profile – |  |
| - Number of B lines | 35 (0-82) |
| - Number of WF signs | 0 (0-8) |
| - Zones with B profile | 9 (5-12) |
| C profile – |  |
| - Number of SPCs | 1 (0-9) |
| - Number of Shred signs | 2 (0-13) |
| - Number of zones with C profiles | 2 (0-13) |
| Total Number of involved areas (B or C profile) | 10 (6-13) |

(WF= waterfall; SPC= subpleural consolidation)

The number of involved and uninvolved areas were significantly different between the three classes of severity (p=0.0026 mild vs moderate, p<0.0001 mild vs severe, p=0.0175 moderate vs severe) and for patients that died vs. those who were discharged (p=0.0249). The correlation between lung ultrasound abnormalities and the SF/PF ratio are summarized in *Table 4*.

**Figure 2.**
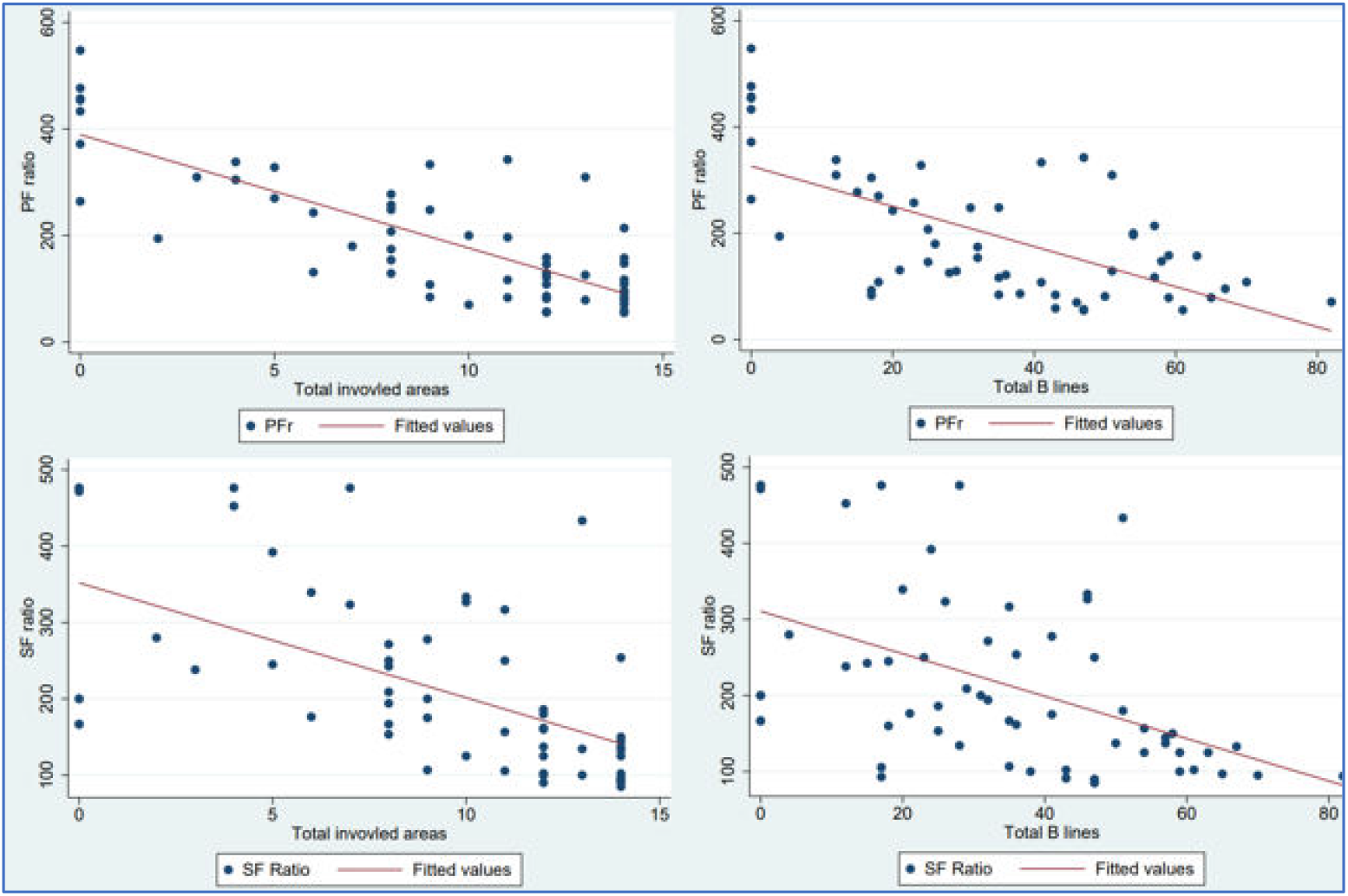
Scatter plots.

**Table 4.** Correlation coefficients.

| SF Ratio | Spearman's $\rho$ | p-value |
| --- | --- | --- |
| Total number of involved areas | -0.6866 | <0.0001 |
| Total number of B lines | -0.5433 | <0.0001 |
| Total number of C profiles | -0.3125 | 0.0151 |
| PF Ratio |  |  |
| Total number of involved areas | -0.7232 | <0.0001 |
| Total number of B lines | -0.5973 | <0.0001 |
| Total number of C profiles | -0.5092 | 0.0001 |

The total number of B lines were predictive of mortality (p=0.0188, OR 1.03, 95% CI 1.003-1.060) and a trend towards significant prediction was seen with no. of involved areas (p=0.0512, OR 1.126, 95% CI 0.993-1.276). After univariate analysis and adjustment for collinearity, the variables used for multivariate regression model for prediction of mortality with the optimized AUC included CRP, anticoagulation use and total no. of B lines (p=0.0124, pseudo R^2^=0.1740), with AUC = 0.7682 (95% CI= 0.6176-0.9188).

The total number of involved areas could differentiate severe from non-severe (mild/moderate) cases (p<0.0001, OR 1.44, 95% CI 1.194-1.743). After univariate analysis and adjustment for collinearity, multivariate regression model with the optimized AUC included LDH and total number of involved areas (p <0.0001, Pseudo R^2^=0.3822) with AUC= 0.8743 (95% CI= 0.775-0.973). A cut-off of ≥6 involved areas (of the 14 areas assessed) was found to have a sensitivity of 100% and specificity of 52% (LR+ = 2.0833) for differentiating severe from non-severe cases.

**Figure 3.**
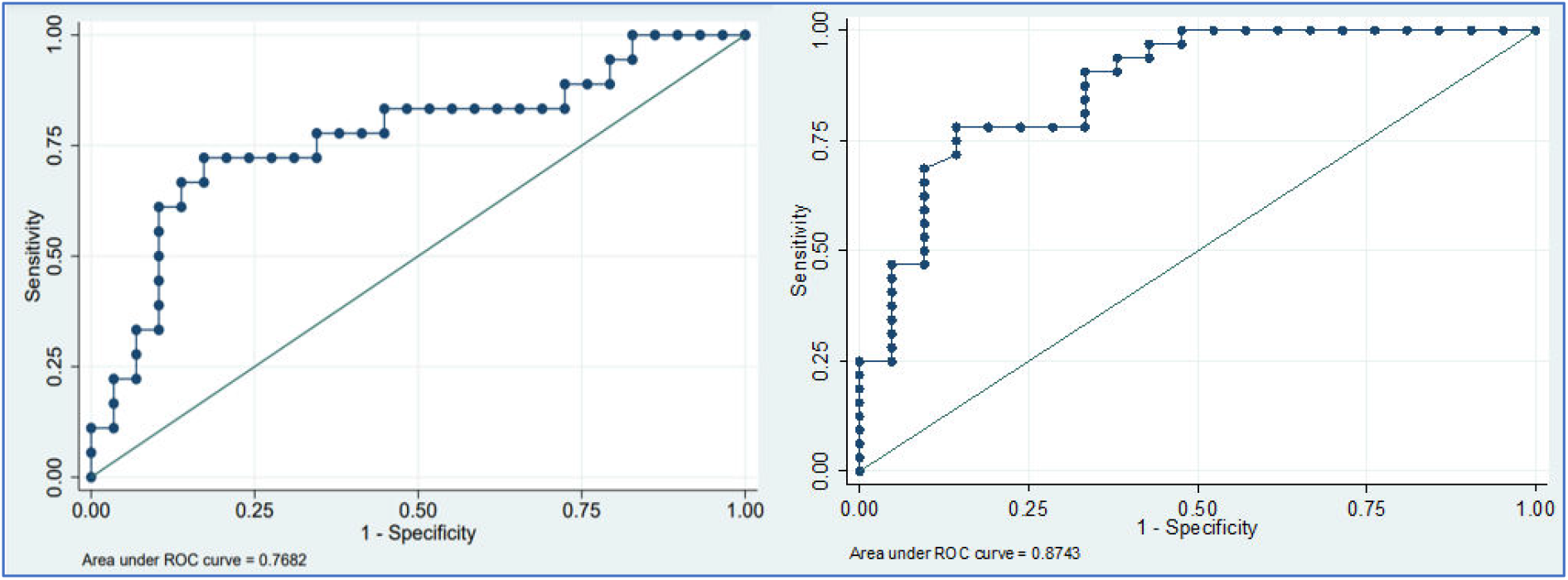
ROC curve for outcome (left) and severity (right) prediction models.

## Discussion

Lung ultrasound is a useful alternative to chest CT or chest X-ray for the diagnosis and follow up of COVID-19 patients^4,8^ but inadequate evidence exists regarding its triage capabilities. The results of this study, which involved a systematic 14-zone lung ultrasound assessment of 60 admitted patients with COVID-19 show that the severity and outcomes correlate with the number of involved areas and the severity of involvement (no. of B lines).

In our study, B–profile was the most common abnormality (88.33% of participants), a result which is consistent with the results from previously published data on lung ultrasound in COVID-19 infection (pooled frequency of B-profile= 0.97, 95% CI = 0.94-1).^4^

We found that the number of involved areas on ultrasound imaging increases with worsening severity and negatively correlates with the PF ratio and SF ratio. This concordance between sonographic involvement and clinical severity is similar to that reported by Lichter et al where higher baseline LUS scores and increase in these scores during hospitalization were associated with severe disease.^9^

Significantly more areas were involved in patients that died vs those that survived. Total number of B lines were predictive of mortality and a model that included number of B-lines, CRP levels and anticoagulation predicted mortality with an optimised AUC=0.7682 (95% CI). While previous studies have noted worse clinical outcomes in patients with higher scores on sonographic involvement ^10^, ours is the first that has found significant prediction of mortality using ultrasound findings.

We devised a model for predicting severe disease (vs mild/moderate severity) that utilized the number of involved areas and LDH (p<0.001, CI 0.775-0.973, AUC= 0.8743). Similar associations between B line density, number of affected zones and clinical severity have been previously reported.^8^ Additionally, number of involved areas even when used alone were a significant predictor of severe disease. In the setting of limited hospital resources strained by the ongoing pandemic, this finding may allow lung ultrasonography to aid in the identification of patients likely to require ICU care.

Apart from the advantages of being a quick, simple, point of care modality with proven correlation with CT severity scores ^11,12^, it also obviates the need for shifting each patient for CT, and provides diagnostic support. Our study further adds to its possible use for triage and prognostication.

Our study is, however, not without limitations. Our study did not include follow-up imaging of the enrolled patients. It was a single centre study with a limited sample size and the inter and intra-observer reliability was not assessed. Scans were performed at variable days after symptom onset, at different stages of the disease. Use of the curvilinear probe rather than a linear probe could have compromised detailed assessment of pleural line abnormalities. Therefore, larger prospective studies using lung ultrasound at different points of patient contact are needed to provide further details on its diagnostic and prognostic performance in different clinical settings in the management of COVID-19 patients.

## Conclusion

Bedside ultrasound is a highly promising, fast and cost-effective diagnostic and likely prognostic modality in COVID-19 patients. Our study proves that the degree of lung involvement on ultrasound correlates with the clinical severity and outcomes, as well as can be used to predict severity and outcomes at the point of first contact, to triage and prognosticate respectively. This association maybe tested further using larger prospective studies to triage and prognosticate COVID-19 patients.

## Possible improvement and future studies

Although a significant association were found with the severity of lung USG findings and the clinical severity and outcomes, the changes in lung ultrasound with the natural course of the disease-improving or worsening, could not be analysed. Hence a longitudinal cohort study reflecting USG involvement changes, that might help us predict a possible worsening in the natural course of the disease, and hence pre-emptively upscale treatment, as well as provide more robust evidence for its use as a diagnostic and prognostic modality for COVID-19 pneumonia.

## Data Availability

All data referred to in the manuscript shall be provided when asked for

## Acknowledgement

CN and SK contributed to study design, data collection, data analysis and interpretation, and writing of the report. MS helped in data collection. PP contributed to data analysis and editing the manuscript. AK, NW, KDS, RA and AT contributed to the study design, and approved the final version of the submitted report.

