## supplement tables and figures for "Clinical correlation of lung ultrasound profiles in patients with COVID-19 infection"

**SUPPLEMENTARY MATERIAL**

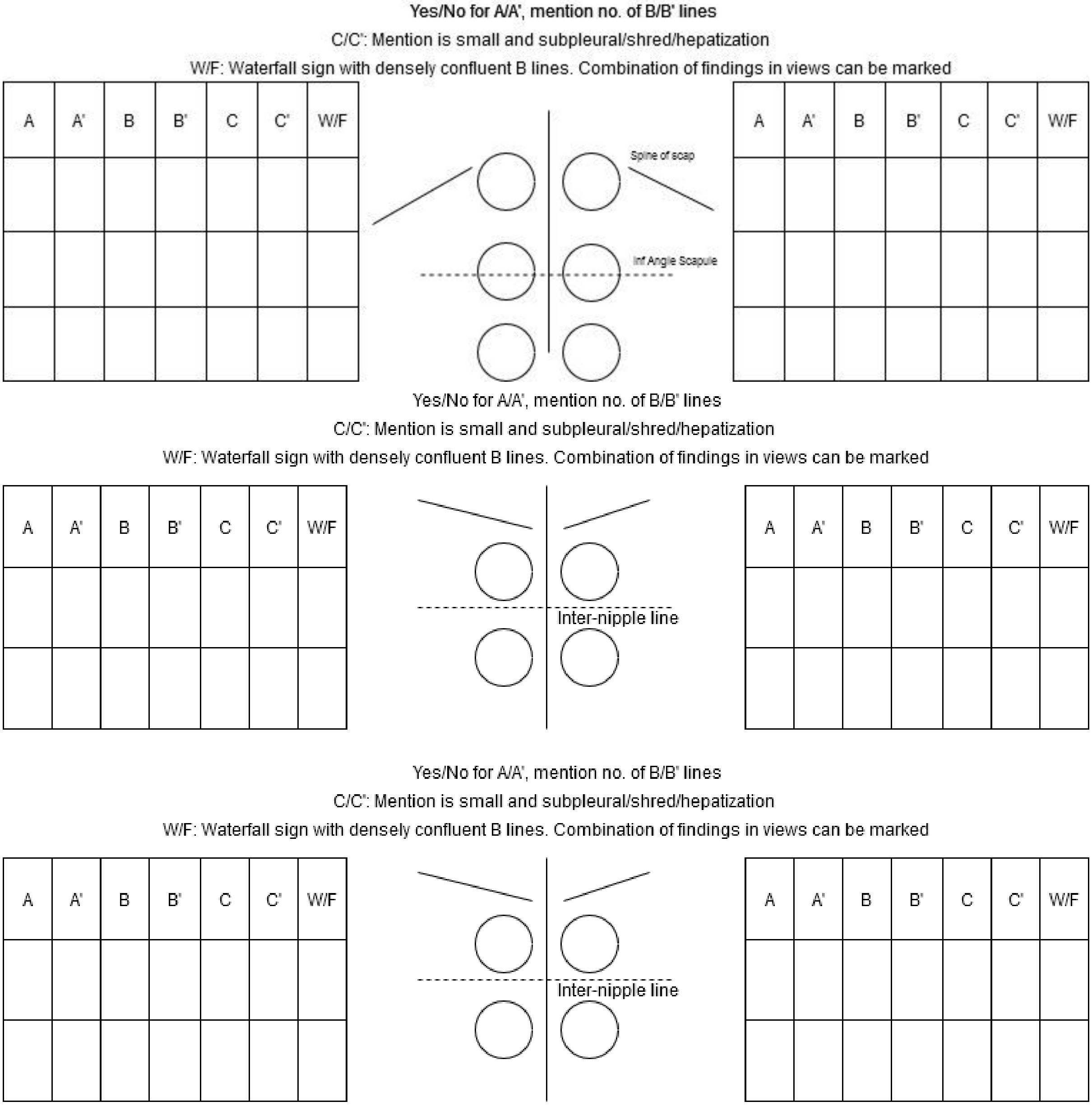


***Figure 1S*. Areas of lung where Ultrasound was performed.**

(Top- Posterior, Middle- Axillary, Bottom- Anterior)

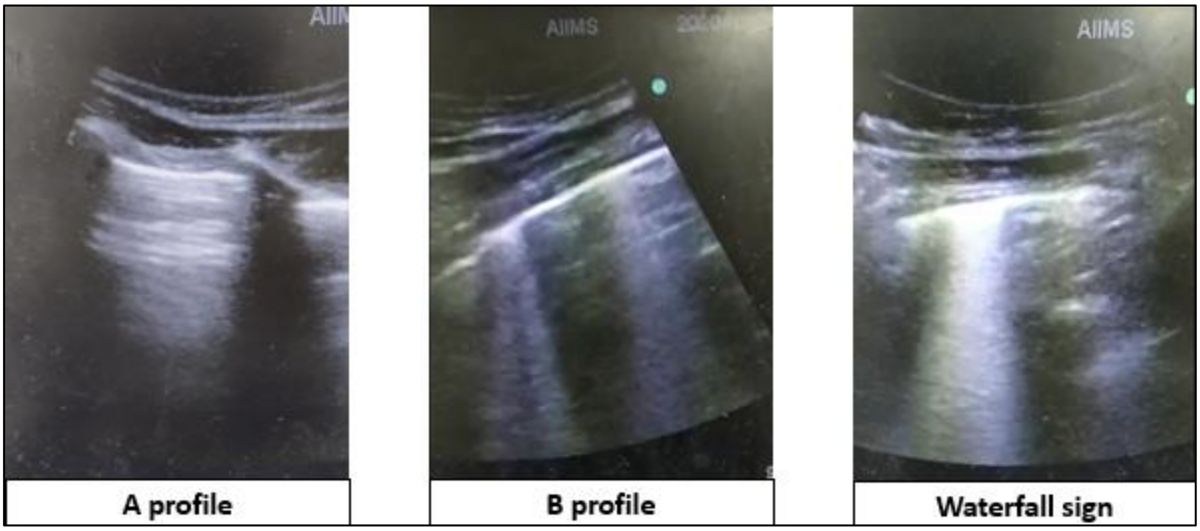


*Figure 2S*. **Lung ultrasound profiles**
